# Embryonic lethal genetic variants and chromosomally normal pregnancy loss

**DOI:** 10.1101/2020.10.16.20214163

**Authors:** Jennie Kline, Badri N. Vardarajan, Avinash Avabhyankar, Sonja Kytömaa, Bruce Levin, Nara Sobreira, Andrew Tang, Amanda Thomas-Wilson, Ruiwei Zhang, Vaidehi Jobanputra

## Abstract

**STUDY QUESTION:** Are rare genetic variants in the conceptus associated with chromosomally normal pregnancy loss?

**SUMMARY ANSWER:** The proportion of probands with at least one rare variant is increased in chromosomally normal loss conceptuses compared with controls.

**WHAT IS ALREADY KNOWN:** Among non-consanguineous families, one study of seven chromosomally normal losses to four couples with recurrent pregnancy loss (RPL) and a case report of a family with RPL of which one was known to be chromosomally normal identify compound heterozygote variants in three different genes as possibly causal. Among consanguineous families, RPL of chromosomally normal pregnancies with non-immune hydrops fetalis (NIHF) has been attributed to recessive variants in genes previously implicated for NIHF and new candidate genes.

**STUDY DESIGN, SIZE, DURATION:** The starting sample was 52 chromosomally normal losses to 50 women, identified in 2003-2005 as part of a cohort study on trisomy and ovarian aging. The analytic sample comprises 19 conceptus-parent trios with DNA from 17 biologic parents (cases). The control group derives from the National Institutes of Mental Health’s National Database for Autism Research (NDAR). It comprises 547 trios of unaffected siblings of autism cases and their parents.

**PARTICIPANTS/MATERIALS, SETTING, METHODS:** We use exome sequencing to identify rare variants in the coding region of the genome. We defined variant rarity in two ways: ultra-rare (absent in gnomAD) and rare (heterozygote <10^−3^ in gnomAD). For autosomal recessives, we further required that the variant was absent as a homozygote in gnomAD. We compare the rates of rare predicted damaging variants (loss of function and missense – damaging) and the proportions of probands with at least one such variant in cases versus controls. Secondarily, 1) we repeat the analysis limiting it to variants in genes considered causal in fetal anomalies and 2) we compare the proportions of cases and controls with damaging variants in genes which we classified as possibly embryonic lethal based on a review which was blinded to case-control status.

**MAIN RESULTS AND THE ROLE OF CHANCE:** The rates of ultra-rare damaging variants (all *de novo*) are 0.21 and 0.17 in case and control probands, respectively. The corresponding rates for rare potentially pathogenic *de novo* variants are 0.37 and 0.24, respectively; for autosomal recessive variants they 0.11 and 0.03. The proportions of probands with at least one rare potentially damaging variant were 36.8% among cases and 22.9% among controls (odds ratio (OR) = 2.0, 95% CI 0.9, 3.0). Secondary analyses show no damaging variants in fetal anomaly genes among case probands; the proportion with variants in possibly embryonic lethal genes was increased in case probands (OR=14.5, 95% CI 3.4, 61.1). Cases had variants in possibly embryonic lethal genes *BAZ1A, FBN2* and *TIMP2. Post hoc* review of these cases suggests that *CDH5* may also be an embryonic lethal gene.

**LIMITATIONS, REASONS FOR CAUTION:** The number of case trios (n=19) limits the precision of our point estimates. We observe a moderate association between rare damaging variants and chromosomally normal loss with a confidence interval that includes unity. A larger sample is needed to estimate the magnitude of the association with precision and to identify the relevant biological pathways.

**WIDER IMPLICATIONS OF THE FINDINGS:** Our data add to a very small literature on this topic. They suggest rare genetic variants in the conceptus may be a cause of chromosomally normal loss.

**TRIAL REGISTRATION NUMBER:** N/A.

**STUDY FUNDING/COMPETING INTERESTS(S):** Exome sequencing of case trios was performed by Baylor-Hopkins Center for Mendelian Genomics through National Human Genome Research Institute grant 5U54HG006542.

Data used in the preparation of this manuscript were obtained from the National Institute of Mental Health (NIMH) Data Archive (NDA). NDA is a collaborative informatics system created by the National Institutes of Health to provide a national resource to support and accelerate research in mental health. Dataset identifier(s): src_subject_id. This manuscript reflects the views of the authors and may not reflect the opinions or views of the NIH or of the Submitters submitting original data to NDA.

No author has a competing interest.

## Introduction

Pregnancy loss <20 weeks gestation (miscarriage), is the most common adverse outcome of pregnancy, occurring in about 30% of pregnancies that survive to implantation and 10-20% of pregnancies that survive to six weeks (Zinaman *et al*., 1996; Wang *et al*., 2003; Wilcox, 2010). Chromosomal abnormality in the conceptus is the cause of at least 50% of losses, with the proportion determined mainly by maternal and developmental (e.g., prefetal, fetal) ages of the sample (Kline *et al*., 1989; Warburton, 2000; Hardy *et al*., 2016). The abnormality usually results in embryonic or fetal lethality (hereafter, embryonic lethality).

The remaining pregnancy losses are chromosomally normal. Aside from an association with maternal age and a few rare conditions, there are few established risk factors for these losses (Kline *et al*., 1989; Cohain *et al*., 2017). It has long been hypothesized that variations in single genes are a cause of chromosomally normal loss. Embryonic lethal variants may arise *de novo* or they may be inherited. We hypothesize that *de novo* variants predominate for three reasons. First, the increased risk of loss after chromosomally normal loss is modest, on the order of 1.4-2 fold (Boue *et al*., 1975; Lauritsen, 1976; Kline *et al*., 1989). Second, among women with repeat losses, the increased risk of a second chromosomally normal loss is modest (Warburton *et al*., 1987). Recent studies suggest a higher recurrence risk for chromosomally normal loss, but do not adjust for maternal age (Sugiura-Ogasawara *et al*., 2012; Nikitina *et al*., 2020). The increased recurrence risk for chromosomally normal loss is detectable among women <35, but not among older women because of the increasing risk of trisomy with age (Warburton, 1987). Third, the morphologic phenotypes are heterogeneous. Most chromosomally normal losses are of pregnancies that did not reach the fetal stage of development (<8 weeks post-conception, hereafter, prefetal). In one embryoscopic study, among 56 chromosomally normal, predominantly prefetal, losses, 29% are classified as normal embryos, 36% as growth disorganized embryos and the remainder with combined or isolated defects (Philipp *et al*., 2003) (our computations). Of the latter group, eight embryos/fetuses (14% of the 56) had structural anomalies, with the remainder involving a phenotype that likely reflects autolysis *in utero* or amniotic adhesion or band anomalies.

Most searches for variants in single genes that disrupt *in utero* development, ordinarily using exome sequencing (ES), focus on structural anomalies, whether diagnosed prenatally or at birth or later. These studies show that genes associated with anomalies among pediatric patients are associated with the same anomalies detected earlier in fetal development (Lord *et al*., 2019; Petrovski *et al*., 2019); see also Best et al (Best *et al*., 2018). Some of these variants may have variable phenotypes, extending to the phenotypes seen in chromosomally normal losses. By analogy with aneuploid losses, however, we hypothesize that chromosomally normal loss is caused by rare variants in many different genes, with some incompatible with development to the fetal stage.

In pregnancy loss, many studies focus on the woman, seeking to identify maternal genotypes associated with recurrent pregnancy loss (RPL). These studies focus on genes related to selected processes (e.g., immune response, metabolism, coagulation, angiogenesis). Findings are inconsistent, with positive studies reporting only modest associations (Rull *et al*., 2012; Pereza *et al*., 2017; Shi *et al*., 2017).

Few studies have sought to identify genetic variants in losses occurring before a diagnosed structural anomaly. A major limitation of most studies is the absence of information on the chromosomal constitution of the conceptus. Pregnancy loss studies have focused on women with RPL, assuming implicitly that repeat losses have the same cause. This assumption is reasonable in consanguineous couples, but less so otherwise. A widely cited study of consanguineous couples with RPL ascertained the sample by the presence of lethal non-immune hydrops fetalis (NIHF) in at least two pregnancies (Shamseldin *et al*., 2015; Shehab *et al*., 2017). Of 19 probands studied, five were chromosomally normal losses of which two were homozygous for genetic variants known to cause NIHF and three for novel candidate embryonic lethal variants. Case reports of consanguineous families with RPL and an index chromosomally normal loss also identify recessive or X-linked variants as causal (Shehab *et al*., 2017). Several case reports or series in non-consanguineous couples that are sometimes classified as RPL studies base ascertainment on recurrence of a structural anomaly in two pregnancies (see Table 2 in review by Colley et al 2019 (Colley *et al*., 2019)). The underlying assumption is that RPL and structural anomalies in the same family have a common cause. It is unclear whether this assumption is valid. Even if it is, findings from these studies will not generalize to most women or couples with RPL, nor with sporadic chromosomally normal loss.

In non-consanguineous couples, repeat losses may have different causes (e.g., a mix of chromosomally normal and abnormal or abnormal of different types). Several studies hypothesize that the maternal (and in one instance paternal) genotype is related to RPL through repeat triploidy (Filges *et al*., 2015) or trisomy (Bolor *et al*., 2009; Mizutani *et al*., 2011; Hanna *et al*., 2012; Sazegari *et al*., 2014). In these studies, although the parental genotype is examined, the hypothesis is that aneuploidy in the conceptus caused the loss. Of the few studies attempting to discover embryonic lethal variants in single genes in RPL, only three (Qiao *et al*., 2016; Fu *et al*., 2018; Mitra *et al*., 2020) draw on conceptuses known to be chromosomally normal.

A study of seven chromosomally normal loss conceptuses (Qiao *et al*., 2016) identifies as potentially causal two compound heterozygote variants in repeat chromosomally normal losses to two couples. A similar finding stems from a case report of a couple with three first trimester losses (Mitra *et al*., 2020), at least one of which was chromosomally normal. These two reports identify two genes that are associated with structural anomalies (*DYNC2H1* and *DHCR7*) suggesting that, in some instances, variants in genes associated with structural anomalies may reflect the lethality of already identified features of the associated syndromes or expansion of the phenotypes to include arrested embryonic development without structural anomalies.

The largest published study includes 19 chromosomally normal loss conceptuses (Fu *et al*., 2018). The investigators assessed the occurrence of damaging SNVs in 286 candidate genes considered related to embryonic lethality. Interpretation is limited by the absence of parental sequencing data (to determine whether variants in the conceptus are *de novo* or inherited), a comparison group, and description of how the genes were selected.

In this paper we add to a small body of literature to estimate the association of rare potentially damaging SNVs with chromosomally normal loss. We draw on an unselected sample in which we obtained DNA from the chromosomally normal conceptus (proband) and parents. We compare the frequency and types of rare potentially damaging SNVs in the 19 trios (cases) with those in 547 unaffected siblings of autism case trios (controls).

## MATERIAL AND METHODS

### Study population

Cases: The sample is drawn from a consecutive series of pregnancy losses <18 weeks developmental age identified at a single hospital (2003-2005) (Warburton *et al*., 2009). The current study includes all chromosomally normal losses for which we had stored conceptus and maternal DNA and the woman consented to use of stored samples in future studies on reproduction.

We submitted 61 conceptus and maternal DNA samples to Baylor Hopkins Center for Mendelian Genetics for ES (Supplement Table S1a sets out the derivation of the sample). Of the 61 samples, nine were excluded from analysis prior to ES (Table S1a, footnote 2). The remaining 52 chromosomally normal losses occurred to 50 couples; two couples each had two chromosomally normal index losses.

Forty-seven (49 losses) women agreed to future contact. In 2016-2018 we attempted to contact these women to update their obstetric histories and to obtain paternal saliva samples (for analysis of DNA). We updated the obstetric histories of 25 women (27 losses) and obtained DNA from saliva from 17 biologic fathers (19 losses).

The primary analytic sample comprises the 19 trios. In addition, we had permission to analyze DNA from all conceptus − mother duos. Of the 33 duos, 28 have good quality ES data (Table S1b and see below). We use these data to define the sample of variants included in the analysis.

We use pathology laboratory data and reported last menstrual period, gestational age and sonography findings to classify losses as prefetal or fetal. The pathology data are of limited utility in part because products of conception were obtained by suction curettage. We define as prefetal, losses that occurred <71 days gestation (from last menstrual period) or in which the conceptus was judged by ultrasound to be non-viable or to have stopped development <71 days. We define as fetal, losses that occurred at ≥71 days and in which ultrasound findings indicated viability at ≥71 days or a fetus or fetal parts were identified in pathology. Among the 19 trios, 12 were prefetal, 5 fetal and 2 of unknown morphology. Mean gestational age was 74 days (range 30-119). At the follow-up interview, mean maternal age was 46 years (range 39-55) and of the 17 women, 13 (76%) reported two or more pregnancy losses (including the index loss).

#### Controls

We selected controls on January 13, 2017 from the National Institutes of Mental Health’s National Database for Autism Research (NDAR) (Hall *et al*., 2012; NIH, 2020). The control sample comprises unaffected siblings of autism cases with exome sequencing performed on DNA from the sibling, mother, and father. We further required the sibling be aged 60-251 months at interview (to limit the chance of including a child with undiagnosed autism). For families with more than one sibling in the NDAR database, we include only one sibling (the oldest at time of interview). In total, 1475 sibling-mother-father trios met these criteria. From these, we selected the trios where the sequencing platform and exome coverage in the sibling and parents was comparable to that used for cases (see below). Of the 1475, 587 families met these criteria. Of these, the genetic sequencing data files turned out to be corrupted (n= 26) or ES data of poor quality (n=14, see below), resulting in a sample of 547 control trios.

### Exome Sequencing

For cases, the proband and maternal exome was captured using the Agilent SureSelect HumanAllExon V4.51 MbKit_S03723314 kit (Agilent v4 capture kit). The paternal exome was captured using the Agilent SureSelectXT Clinical Research Exome. For controls, the exome was captured using NimbleGen custom array or EZExomeV2.0(Sanders *et al*., 2012).

#### Variant-calling

We used New York Genome Center’s automated analysis pipeline to jointly call variants in cases and controls (Arora *et al*., 2019). We aligned sequencing reads to the human reference GRCh37 using BWA-MEM v0.7.15 (DePristo *et al*., 2011; Van der Auwera *et al*., 2013). We called and recalibrated variants using the GATK best-practices.

#### Exclusions

We used the PLINK’s genome-wide IBD sharing algorithm (Purcell *et al*., 2007; Chang *et al*., 2015) to check for relationships within and between pedigrees and homozygosity of the X-chromosome to compare phenotypic with genotypic sex. We exclude families that fail either the relationship or sex checks. Among cases, we exclude five conceptus − mother duos (Table S1b). Among controls, we exclude 14 trios.

#### Case-control comparability

The exome capture library kits used in cases capture 51 MB of the genome, while the kit for controls captures 37 MB of the genome. To avoid bias in detecting putatively causal variants, we limit our analysis to the intersection of the two capture kits such that each qualifying variant was called (as homozygous reference or non-reference) in at least 80% of 123 case samples (includes 19 trios and 28 conceptus − mother duos) and 80% of 1641 control samples (547 trios). This eliminates variants that occur in off-target regions or regions covered unevenly between cases and controls.

#### Primary analysis

We define as damaging variants with predicted loss of function (i.e., nonsense, splice site, and frameshift, hereafter, LoF) and missense variants with REVEL>0.50 (Ioannidis *et al*., 2016). For *de novo* and X-linked variants we carry out the analysis twice, first using a strict rarity criterion (i.e., the variant in the proband did not occur in gnomAD (Karczewski *et al*., 2020), hereafter “ultra-rare”) and second, relaxing the rarity criterion (i.e., overall minor allele frequency (MAF) in gnomAD <10^−3^, “rare”). For homozygous and compound heterozygote (CH) variants, we also required that the variant be absent as a homozygote in gnomAD. We hypothesize that embryonic lethal variants are incompatible with survival to birth and normal development. Thus, in our analyses, we exclude *de novo* variants in probands that are also observed in another unrelated member of the sample (parents or probands), excepting variants that occur in more than one proband in the same case or control group.

We compare cases and controls for: 1) the rates of variants of specific types (LoF, missense – damaging, missense – tolerated and synonymous) and mode of inheritance and 2) the proportion of probands with at least one damaging (LoF or missense – damaging) variant. We use synonymous variants to determine whether case and control data are comparable. We test the significance of differences in case-control differences in rate ratios with a conditional two-tailed binomial test and the case-control differences in the proportions with a two-tailed Fisher’s exact test. We use *P*<0.05 as the threshold in testing the statistical significance of each comparison and provide 95% confidence intervals for the odds ratios (ORs) in the second comparison. We do not adjust for multiple comparisons. We provide data for cases and controls in the supplement (Supplement Tables S2-S5) to allow others to test or generate hypotheses.

Secondary analyses: First, we repeat the analysis using the rare criteria, limiting the analysis to 1340 genes classified as causing or likely to cause fetal anomalies and taking account of allele type associated with disease (Lord *et al*., 2019). Second, we compare the frequency of damaging variants that occurred in genes likely to be lethal to the embryo. To classify genes and variants as suspect, two investigators (AT and VJ) reviewed the literature on all genes that occur in damaging variants in our sample without knowledge of case-control status. In PubMed, they searched for “GeneX NOT cancer NOT tumor NOT carcinoma”. Depending on the initial search results, the added terms including pregnancy or miscarriage or abortion or stillbirth or development. For genes without a well-established association with human disease or for which the initial search yielded very little information, they expanded the search to animal or *in vitro* models. They evaluated the variants based on the guidelines of the American College of Medical Genetics and Genomics (Richards *et al*., 2015) under the assumption that the variant occurred in a case proband. Since we were for the most part evaluating genes, rather than specific variants, they did not use the five variant classification categories of the College. Rather, they classified variants in four categories based on what is known about the gene. The categories are: known lethal (there are none), “likely” to be lethal (none) to the embryo, “possibly” lethal or “not likely”. Of the 154 genes (159 variants) in which damaging variants occurred, they classified 10 as possibly lethal (Supplement S6).

## Results

Cases and controls have similar rates of synonymous missense variants. Cases and controls do not differ significantly in either the rates of LoF and damaging *de novo* or recessive, including compound heterozygous (CH), variants nor in the proportion of probands with at least one damaging variant, whether we define variants as ultra-rare or rare (Table I). Among probands, 36.8% of cases had at least one rare damaging variant compared with 22.9% of controls (OR = 2.0, 95% CI 0.9, 3.0).

**Table I.** Genetic variants among 19 chromosomally normal losses (cases) and 547 unaffected siblings of children with autism (controls): a-c) Rate ratios for variants classified by rarity, mode of inheritance and type and d) Odds ratio relating presence of any potentially damaging variant to chromosomally normal loss

|  | Ultra-rare variants<sup>1</sup> |  |  |  | Rare variants<sup>2</sup> |  |  |  |
| --- | --- | --- | --- | --- | --- | --- | --- | --- |
|  | Cases | Controls | Rate ratio | *P*<sup>3</sup> | Cases | Controls | Rate ratio | *P* |
| **a. *De novo* (rate, number)<sup>4</sup>** |  |  |  |  |  |  |  |  |
| Synonymous | 0.26 (5) | 0.23 (127) | 1.13 | 0.28 | 0.52 (10) | 0.47 (259) | 1.11 | 0.30 |
| Missense – Tolerated | 0.63 (12) | 0.54 (296) | 1.17 | 0.24 | 0.84 (16) | 0.93 (511) | 0.90 | 0.60 |
| Missense – Damaging | 0.16 (3) | 0.14 (76) | 1.14 | 0.27 | 0.26 (5) | 0.19 (104) | 1.38 | 0.16 |
| Loss of function | 0.05 (1) | 0.03 (17) | 1.69 | 0.12 | 0.11 (2) | 0.05 (25) | 2.30 | 0.06 |
| Any damaging | 0.21 (4) | 0.17 (93) | 1.24 | 0.23 | 0.37 (7) | 0.24 (129) | 1.56 | 0.09 |
| **b. Recessive-homozygous (rate, number)** |  |  |  |  |  |  |  |  |
| Synonymous |  |  |  |  | 0.00 (0) | 0.03 (14) | 0.00 | 0.38 |
| Missense – Tolerated |  |  |  |  | 0.00 (0) | 0.03 (14)<sup>5</sup> | 0.00 | 0.38 |
| Missense – Damaging |  |  |  |  | 0.00 (0) | 0.004 (2) | 0.00 | 0.07 |
| Loss of function |  |  |  |  | 0.00 (0) | 0.00 (0) | NA | NA |
| Any damaging |  |  |  |  | 0.00 (0) | 0.004 (2) | 0.00 | 0.07 |
| **c. Recessive-compound heterozygous (rate, number)** |  |  |  |  |  |  |  |  |
| Both synonymous | NA | NA |  |  | 0.26 (5) | 0.25 (138) | 1.04 | 0.35 |
| Any damaging<sup>6</sup> | NA | NA |  |  | 0.11 (2) | 0.03 (19) | 3.03 | 0.03 |
| **d. Probands with at least one damaging variant (percent, number)<sup>7</sup>** | NA | NA |  |  | 36.8 (7) | 22.9 (125) | 1.97 | 0.17 |

<sup>1</sup> For both *de novo* and recessive modes, we define as “ultra-rare” variants that are not present in gnomAD.
<sup>2</sup> We define “rare” variants as either absent in gnomAD or present as heterozygotes with MAF<0.001 in gnomAD. For recessive homozygous and compound heterozygote (CH) variants, we require that neither variant occur as a homozygote in gnomAD.
<sup>3</sup> We test the significance of the case-control differences in the rate ratios with a conditional two-tailed binomial test. We test the significance of the odds ratio with a two-tailed Fisher’s exact test.
<sup>4</sup> Missense – tolerated = missense variants with REVEL $\leq 0.5$ (Ioannidis *et al.*, 2016); Missense – Damaging = missense variants with REVEL $> 0.5$ ; Loss of function (LoF) variants include nonsense, splice site, and frameshift variants. Any damaging = Damaging missense + LoF.
<sup>5</sup> Includes two male probands with X-linked recessive variants.
<sup>6</sup> For CH, we define as damaging CH in which both variants are damaging (Missense – damaging or pLoF). We show the rates of CH with two synonymous variants to indicate the comparability of case-control data. The potentially damaging CH variants included four LoF variants (1 in cases, 3 in controls).
<sup>7</sup> We use the “rare” criteria to identify the numbers of cases and controls with at least one damaging variant.

Secondary analyses: No cases and 15 controls have at least one damaging variant in a gene associated with fetal anomalies. Among controls, the damaging variants occur in 13 different genes (Supplement S2, S4).

Three cases and seven controls have at least one damaging variant in a gene classified as “possibly” embryonic lethal (OR = 14.5, 95% CI 3.4, 61.1, *P*=0.003). Among cases, two variants, in *BAZ1A* and *TIMP2A*, are *de novo*, and one, in *FBN2*, is a compound heterozygote (Supplement S7).

One case had a *de novo* nonsense variant in *BAZ1A*, c.[1156C>T]; p.[Arg386Ter]. BAZ1A (Bromodomain Adjacent to Zinc Finger Domain, 1A; MIM#605680) is involved in chromatin remodeling. The variant has a pLi of 1 (o/e 0.1; 8 observed, 82 expected in gnomAD). While *BAZ1A* does not currently have an OMIM gene-disease association, two reports describe *BAZ1A* missense variants in individuals with multisystemic syndromic phenotypes (Zaghlool *et al*., 2016),(Weitensteiner *et al*., 2018). Thus, we speculate that a nonsense variant in *BAZ1A* might disrupt embryonic development. This same case had a *de novo* missense variant in *RRAD* (Supplement S8), however, given that there are >50 heterozygous individuals in gnomAD, we think it unlikely that this variant is related to pregnancy loss.

One case had a *de novo* damaging missense *TIMP2* variant, c.[553G>A]; p.[Gly185Arg]. TIMP2 (Tissue Inhibitor of Metalloproteinase 2; MIM#188825) is an inhibitor of Matrix Metalloproteinase 2 (MMP2; MIM#120360), which is a type IV collagenase. The MMP2/TIMP2 complex is involved in several gestational processes including implantation and placentation (Staun-RamShalev, 2005). We hypothesize that perturbation of these processes may be the cause of the pregnancy loss.

One case had two damaging missense variants in *FBN2*, c.[4457G>T];[2927G>T] and p.[Ser1486Ile];[Gly976Val]. Autosomal dominant variants in *FBN2* (Fibrillin 2; MIM#612570) are associated with Congenital Contractural Arachnodactyly (Beals syndrome, CCA; MIM#121050), which includes skeletal, joint, and muscular manifestations (Callewaert, 1993 updated 2019; Meerschaut *et al*., 2020). Both severe and mild phenotypes have been observed, and inter- and intrafamilial variability is recognized (Callewaert, 1993 updated 2019). In our case, we did not clinically examine the parents and thus do not know whether they manifest a mild unrecognized form of CCA. Since pathogenic variants in *FBN2* can result in fetal akinesia, it is possible that biallelic pathogenic variants in this gene may disrupt embryonic development.

This same case proband had a *de novo* damaging missense variant in *CDH5* (Supplement S8). We did not classify this gene as possibly embryonic lethal *a priori* in part because we accidently missed relevant literature from mouse models. *CDH5* (Cadherin 5; MIM#601120) does not currently have a gene-disease association in OMIM. The c.535G>A (p.Asp179Asn) variant identified in the conceptus is absent in gnomAD, although several variants within the same region occur at low frequencies. *CDH5* is involved in cell differentiation and vascular endothelial development. On discovering that the *CDH5* variant occurred in a case, we re-examined the literature. In homozygous mouse knockout models, *CDH5* is an embryonic lethal; absence is associated with defects in endothelial cell organization and angiogenesis (Gory-Faure *et al*., 1999; Crosby *et al*., 2005). Thus, the one case has *de novo* damaging missense variants in two genes, *FBN2* and *CDH5*, each of which might disrupt embryonic development.

Of the seven controls with at least one variant in a possibly embryonic lethal gene, two had another damaging variant (one in *SLC4A1* and the other in *WDR19*). Both genes are classified as unlikely to be embryonic lethal (Supplement S9).

We studied two chromosomally normal losses to two couples in the sample. In one family, neither conceptus had a potentially damaging variant. In the other family, one conceptus had a potentially damaging variant and one did not.

## Discussion

Our data provide support for the hypothesis that rare genetic variants in the conceptus may cause pregnancy loss. Our sample, however, was too small to estimate the strength of association with precision. Thus, we observed a moderate (OR=2.0), not statistically significant, increase in the proportion of cases with at least one rare potentially pathogenic variant compared with controls. In support of the inference that rare variants are a cause of some chromosomally normal losses, the point estimate for the rate ratios for *de novo* LoF variants is stronger than that for missense – damaging variants (2.3 versus 1.6).

When we limited the analysis to genes associated with fetal anomalies (Lord *et al*., 2019), no case proband has a damaging variant compared with 15 (2.7%) of control probands. Among controls, damaging variants occurred in 13 genes.

One limitation of our study is that we did not assess conceptuses for potentially pathogenic copy number variants. We do not consider this a significant limitation because these variants are rare in chromosomally normal loss conceptus (about 5%) (Zhou *et al*., 2016).

*A priori*, among potentially damaging variants, we classified 10 genes as possibly lethal *in utero*. Variants in possibly lethal genes occurred in three cases and seven controls (OR = 14.5). Among cases, the possibly lethal genes are *BAZ1A, FBN2* and *TIMP2*. If the associations with variants in *BAZ1A* and *FBN2* are causal, we hypothesize a mechanism related to disruption of embryonic development. For *TIMP2* the literature favors mechanisms that interfere with implantation or placentation. Of the remaining 16 cases, four had damaging variants in genes that we did not classify as a possibly embryonic lethal. These include cases with *de novo* damaging variants in *ARSH, FAM131B* and *MORN5* and a compound heterozygous variant in *SERGEF. Post hoc*, in a review of the three case probands with possibly embryonic lethal genes, we discovered that one also had a variant in *CDH5*. Although we now think that *CDH5* is a possibly embryonic lethal gene, we do not include this gene in our case-control comparison. Because the *CDH5* variant occurred in a case already classified as having a variant in a possibly embryonic lethal gene, inclusion would not alter our estimate of the strength of association.

*FAM131B* and *MORN5* are not currently listed in OMIM; their protein function is unknown; we did not find any reports of associations with human or mouse phenotypes. *ARSH* (Arylsulfatase H; MIM#300586) and *SERGEF* (Secretion-Regulating Guanine Nucleotide Exchange Factor; MIM#606051) are listed in OMIM, but without a disease association; we did not find any evidence of associations with human or mouse phenotypes. Current knowledge is insufficient to speculate on a role for any of these genes in pregnancy loss.

Our analyses have several strengths. First, our data from controls—unaffected siblings of autism probands—are consistent with those reported in another similar control sample, supporting the validity of our analysis. In a study of congenital heart disease (Jin *et al*., 2017), at least one potentially damaging *de novo* or autosomal recessive variant was detected in 26% of unaffected siblings of autism proband controls. In our sample, the corresponding rate is 23%. Second, we classified the genes in which damaging variants occurred as possibly embryonic lethal or not without knowledge of case-control status. Thus, unlike prior observations in this area, our comparison is not vulnerable to unintended biases that accompany knowledge of the pregnancy outcome. (Our reconsideration of *CDH5* is an exception to this statement since it is done with knowledge of case-control status.) Third, our sample is larger than any other reported study of chromosomally normal loss conceptus trios thus adding to a small body of literature on such losses in a non-consanguineous sample. On the other hand, our data comprise only 19 trios, limiting not only our ability to make a precise estimate of the strength of associations, but also to identify likely causal variants. Since we hypothesize that many genetic variants contribute to chromosomally normal loss, we did not expect to find damaging variants in the same gene in more than one case in our sample.

In sum, our data suggest that the odds of a lethal variant might be increased by as much as two-fold in chromosomally normal loss conceptus compared with controls. This result requires replication in a larger sample both to confirm the point estimate and to identify biological processes in chromosomally normal loss.

## Supporting information

Supplementary Material Table of Contents

## Data Availability

All raw and processed de-identified data will be shared upon request, as per the consent specified in our IRB approval.

## ACKNOWLEDGEMENTS

We acknowledge the late Dorothy Warburton who conceived this study with us. We acknowledge Judy Chih-Yu and Vanessa Felice who prepared the DNA samples, Amelia Zuver and Troy Layouni who set up the fieldwork databases, and Sarah Robbins who collaborated in the planning stage of the analysis. We thank Kathy Hardy for her insightful feedback on this paper. We thank the women and men who participated in this research.

