## Supplementary Material Table of Contents for "Embryonic lethal genetic variants and chromosomally normal pregnancy loss"

S1	Derivation of the case sample

S2	List of damaging de novo variants

S3	List of damaging recessive homozygous variants

S4	List of damaging recessive compound heterozygote (CH) variants

S5	List of probands with number and type of damaging variant

S6	Classification of genes containing damaging variants as possibly embryonic lethal or not likely

S7	Cases and controls with possible embryonic lethal variants

S8	Damaging variants in seven chromosomally normal loss conceptuses with a variant in at least one possibly embryonic lethal gene

S9	Damaging variants in seven controls with a variant in at least one possibly embryonic lethal gene

S10	List of NDAR controls in the analytic sample: IDs and subject keys
