## Supplementary Table S1 for "Embryonic lethal genetic variants and chromosomally normal pregnancy loss"

Table S1a. Derivation of the sample of chromosomally normal miscarriages for exome sequencing (ES) and the follow-up study (2016-2018)

|  | Number | Number of couples^[1]^ |
| --- | --- | --- |
| Chr. normal miscarriage^[2]^ | 61 | 59 |
| Eligible for ES of miscarriage and woman^[3]^ | 52 | 50 |
| Eligible for follow-up^[4]^ | 49 | 47 |
| Refused follow-up study^[5]^ | 8 | 8 |
| Not located or deceased^[6]^ | 14 | 14 |
| Woman completes follow-up interview | 8 | 8 |
| Woman and man complete follow-up study | 19 | 17 |

Table S1b. Derivation of the sample of chromosomally normal miscarriage trios and duos with usable whole exome sequencing (ES) data

|  | Miscarriage-Woman-Man trios | Miscarriage-Woman |
| --- | --- | --- |
| DNA sequenced | 19 | 33 |
| ES data good quality^[7]^ | 19 | 28 |

ES=Exome sequencing.

1. Two couples each had two chromosomally normal miscarriages.
2. Miscarriage samples chromosomally normal based on tissue culture or FISH with probes for chromosomes 13, 15, 16, 18, 21, 22, X and Y for specimens in which the culture was not successful or yielded a chromosomally normal female karyotype.
3. Baylor-Hopkins Center for Mendelian Genetics screened miscarriage and maternal DNA prior to ES. Nine samples are not eligible for ES: in four, microarray analysis revealed a chromosomal abnormality and in five, the DNA of the miscarriage was not of high enough quality for sequencing. The newly discovered chromosomal abnormalities include trisomy 8 in samples diagnosed with FISH, but not tissue culture (n=2), uniparental disomy of chromosome 1 (not detectable by karyotyping or FISH) (n=1) and a low frequency trisomy 22 mosaic missed by FISH (n=1).
4. At the time of the initial study, three women declined contact about future research. For the remainder we attempted to contact and interview the woman and to contact her partner (i.e., father of the miscarriage) to obtain saliva for DNA and an interview.
5. Refusals include active refusals (n=4) and passive refusals (i.e., we spoke with the woman but she stopped answering our telephone calls) (n=4).
6. Telephone number and address not confirmed (n=12), telephone number but not address confirmed (n=1), deceased (n=1).
7. In four cases, the karyotypic sex of the miscarriage is male and the genotypic sex is female, including two cases where miscarriage and mother were identical. In one case, the miscarriage is related genetically to too many other subjects in the case-control sample.
